# Markovian Random Walk Modeling and Visualization of the Epidemic Spread of COVID-19

**DOI:** 10.1101/2020.04.12.20062927

**Authors:** Haluk Akay, George Barbastathis

## Abstract

The epidemic spread of CoVID-19 has resulted in confirmed cases of viral respiratory illness in more than 1.4 million people around the world as of April 7^th^, 2020 [1]. However, different regions have experienced the spread of this disease differently. Here, we develop a Markovian random-walk spatial extension of a quarantine-enhanced SIR model to measure, visualize and forecast the effect of susceptible population density, testing rate, and social distancing and quarantine policies on epidemic spreading. The model is used to simulate the spread of CoVID-19 in the regions of Hubei, China; South Korea; Iran; and Spain. The model allows for evaluating the results of different policies both quantitatively and visually as means of better understanding and controlling the spread of the disease.

## 1 Introduction

In late 2019, the infectious disease CoVID-19 began spreading globally, from its origin in Wuhan, China, to every populated continent on earth [2] resulting in a pandemic declared officially by the World Health Organization by March 2020 [3]. By April 7^th^, 2020, the death toll stood at 81,865, with 1,426,096 cases confirmed globally [1]. Due to the long time delay between exposure and experience of characteristic symptoms, the apparently common occurrence of asymptomatic yet infectious individuals, and lack of any antiviral treatment during spreading [4], the disease spread rapidly before most regional governments had the time to implement appropriate containment policies.

Even though the viruses exhibit substantial genetic similarity across regions, the nature of its epidemic spread is unique to each region. Of particular economic importance, and its indirect effects on human welfare during the pandemic, is the surge of demand [5] for staple goods at the peak of active confirmed infected cases in a population. This work seeks to understand why different countries experience this peak at different magnitudes and different speeds. Given the contemporary nature of this pandemic, in addition to the aim of purely modeling the epidemic spread of CoVID-19, this work seeks to graphically visualize this spread for the benefit of all concurrent research continuing on this topic. The key information the visualization presented seeks to provide is the temporal and spatial quality of epidemic spreading, given a set of initial conditions and disease spread and containment specifics in the region under study.

The model presented is inspired by temporal visualizations of the spread of forest fires, where a single flaming tree among a lattice of trees can “infect” other “susceptible” green trees with the fire, until every tree has “recovered” by burning down [6]. Each of these actions occur with fixed probabilities that govern the Markovian system; by associating colors with each state, spreading of the disease or “forest fire” may be visualized. Our model adds a spatial component where agents make random walks to mimic how the disease spreads dynamically through human movement.

In order to simulate epidemic spreading, the possible states an agent can assume are based on the Susceptible – Infected – Removed (SIR) model, a set of nonlinear ordinary differential equations which can be used to track the magnitude of each population, assuming a closed system where *S*(*t*) + *I*(*t*) + *R*(*t*) = *N* is constant. In this system, the coefficient *β* represents the rate of transmission of the disease from the infected to the susceptible population, and *γ* represents the rate of removal, corresponding to recovery or death. It is justifiable to lump the latter two populations into one for the purpose of modeling, as long as both cease to be capable of further infecting others. The coupled ordinary differential equations describing the SIR model are

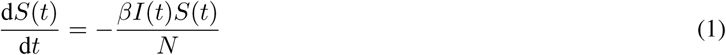

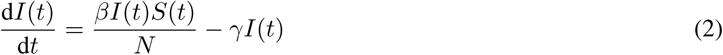

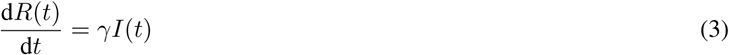

The SIR model has been applied to model the previous SARS coronavirus epidemic [7] [8]. Variations of the SIR model include the SEIR model where a new state *E* (exposed) is introduced, or SIS, where recovered agents are susceptible to reinfection. While these models have been shown to act as accurate approximations for the spread of notable infectious diseases [9], the pandemic nature of CoVID-19 has resulted in policy-driven responses from communities to actively curb spread of the disease, namely quarantining large numbers of individuals testing positive for the virus [10]. This work accounts for this population by extending the model to add a Quarantined state *Q* that captures the effect of isolating agents who test positive for the virus. By applying the SIR-Q states to a spatio-temporal random walk model, the spread of CoVID-19 can be visualized and the effect of critical demographic and policy variables upon epidemic spreading are evaluated.

## 2 Markov Random Walk Model

In order to estimate the spread of CoVID-19 in a human population, a Random Walk framework is used to model transmission of and recovery from the virus in the temporal and spatial domains and provide a visualization of this process. This stochastic process is Markovian in that each subsequent state of the system depends only on the previous state.

### 2.1 Model Framework

The spatial domain is defined as a square 2-dimensional lattice, where agents can exist at discrete nodes. The agents have the ability to make a random walk in one direction at each time-step. The distance of walk is drawn randomly from a Gaussian distribution 𝒩(*µ, σ*^2^) where the mean *µ* is the distance between the starting node and the center of the lattice, and the standard deviation *σ* is one fifth the length of one dimension of the square lattice. At any given time-step, this creates an expectation of a denser population nearer to the center of the lattice.

The model is built on a recursive, Markovian algorithm, where a population of agents distributed over the lattice is initialized, each agent takes a random walk along a single dimension (selected randomly), and finally each agent has the opportunity to change states. A census of the new populations and locations of each agent is taken, and the cycle continues recursively. The simulation terminates when a predefined maximum number of iterations is reached.

Agents are designated as susceptible (S), infected (I), quarantined (Q), or removed (R). The transition of an agent’s status from one state to another is entirely probabilistic and Markovian. The probabilities which govern the model are based on the rates of transmission and recovery defined by the SIR-Q model previously introduced. These rules are listed below and are applied at every time-step.

- If susceptible agents occupy the same node as at least one infected agent, then each susceptible agent may transition to becoming an infected agent with probability *β*. If there are multiple infected agents, then the probability of not becoming infected halves with each additional agent.
- Any agent may be tested with a probability *P*_*t*_. If an infected agent is tested, then they transition to being quarantined, meaning they cannot make movements nor infect other susceptible agents until they recover.
- Any quarantined or infected agent may recover or pass away with probability *γ*, effectively becoming removed from the system.

The spread of disease through the population is visualized by assigning different colors to represent agents of each status occupying each node in the lattice. A frame is taken as a record of the systems state at every time-step. The frames can be compiled into an animation to graphically visualize the spreading, and compare visually how different parameters affect the system.

## 2.2 Parameter Selection and Initial Conditions

The lattice is defined as a 2-dimensional structure, of length 100 nodes in each direction. At initialization, *S*_0_ initial number of susceptible agents and *I*_0_ initial number of infected agents are randomly distributed throughout the lattice. For all the simulations shown in this work, *I*_0_ is initialized as 10 agents. The population density *D* is linearly proportional to *S*_0_, and for simplification purposes, a density index *D* of 1.0 indicates *S*_0_ initialized as 10,000 agents in the following simulations. The testing rate *T* directly corresponds to the probability of an agent being tested *P*_*t*_.

The most critical metric for measuring the spread of a disease is its reproduction number, which is a ratio of the transmission to recovery rate, and a strong indicator of rate of spreading, shown in equation 4.

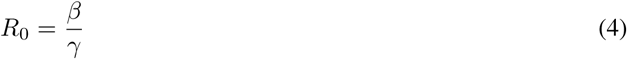

There is no consensus currently on the exact reproduction number of CoVID-19 yet [11], but a study using the SEIR model to estimate *R*_0_ using data from Wuhan, China concluded it fell within a 95% confidence interval of between 5.7 – 7.2, estimated to be 6.5 [12]. Neural-network based models have suggested how effective reproductive number can even be a dynamic variable [13]. For this work, *R*_0_ was taken to be 7.0.

Each calendar day is approximated by one time-step, and a linear population scaling factor was used to fit the model to available data [14] through April 6th 2020 for the Hubei province of China where the city of Wuhan is located, South Korea, Iran, and Spain. The population density indexes *D* and testing rates *T* for each country or region were learned from the data.

## 3 Simulation Results

### 3.1 Effect of Susceptible Population Density and Testing

First, a theoretical environment is considered to measure the effect that changes in susceptible population density and testing rates have on the spread of CoVID-19 in a closed system. While holding the population density index *D* constant at 1.0, the testing rate *T* was altered, and the changes in population of susceptible, infected and quarantined, and removed agents over time were simulated, shown in figure 1. Frames from the visualization of simulation taken at timesteps of 5, 25, 45, and 65, for the system with no testing policy and the system with a testing rate *T* of 0.2 are displayed in figure 3.

**Figure 1:**
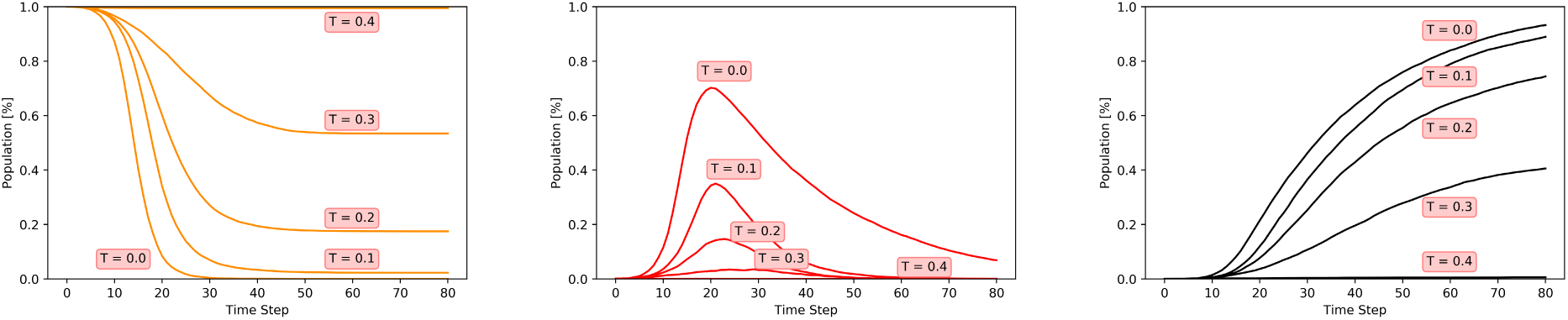
Effect of Testing Rate *T* on S (left), I (middle), R (right) populations.

**Figure 3:**
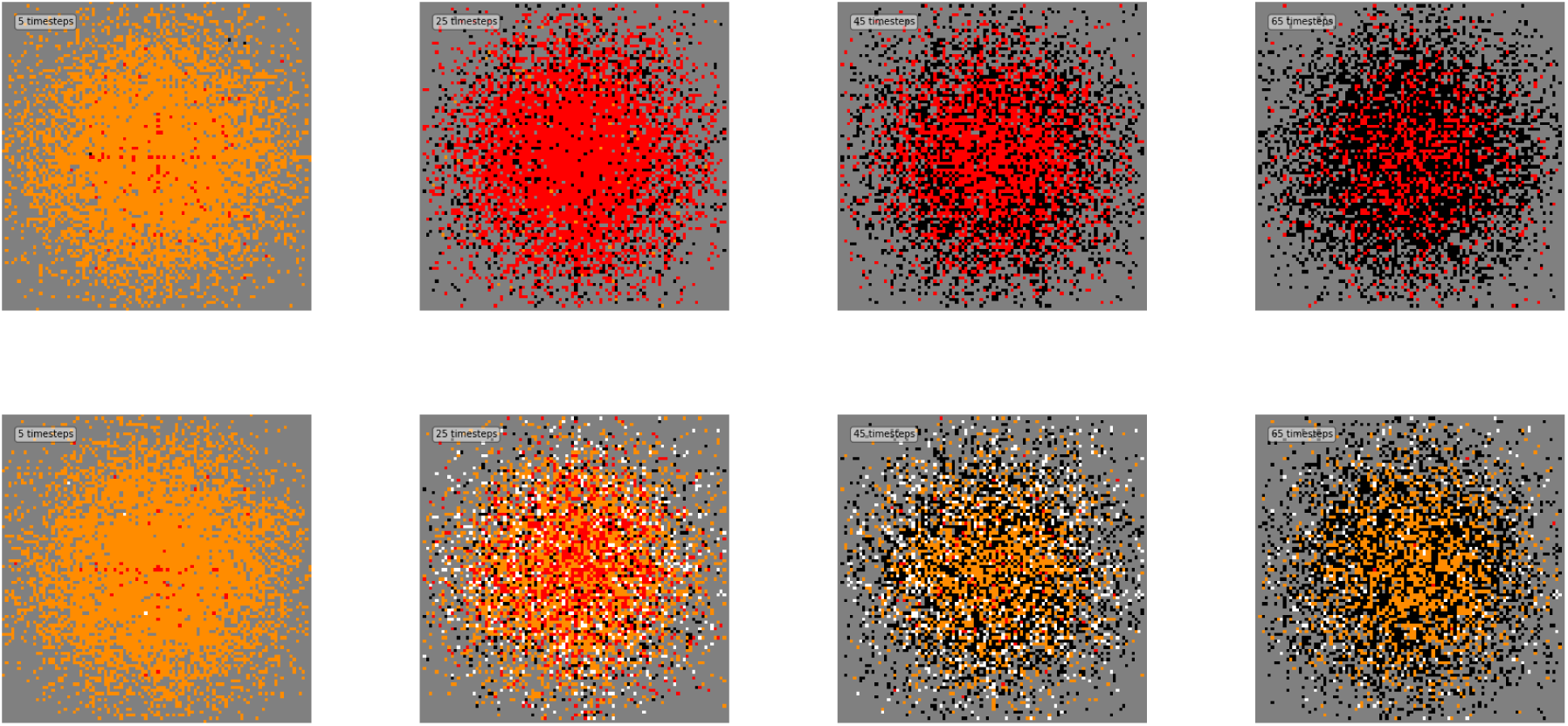
Visualization of effect of instituting Testing Rate *T* of 0.2 (bottom row), compared to no testing (top row). S denoted as yellow, I as red, R as black, Q as white. Time progression from left to right, frames taken at timesteps of [5, 25, 45, 65].

The theoretical environment was also used to demonstrate the effect changes in population density had on the changes in makeup of population with respect to state of all agents in the system. The results of these simulations are shown in figure 2. In both figures 1 and 2, the vertical axes denotes the percentage of total agent population in each closed system.

**Figure 2:**
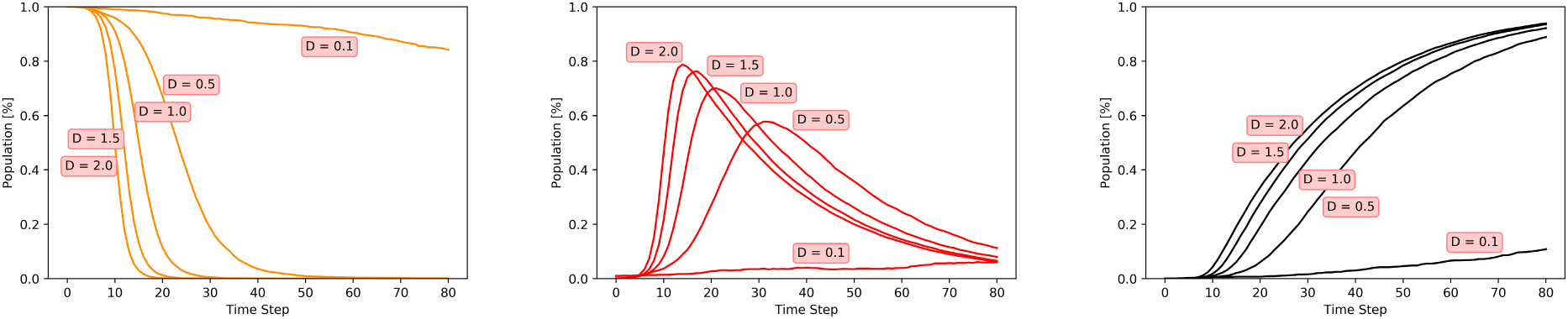
Effect of Population Density on S (left), I (middle), R (right) populations.

The effect of both differing testing rates *T* and population density indexes *D* on the system can be estimated by using the metric of peak infected percentage of the population as a measure of the disease’s maximum impact to the system. The results of these simulations are shown in figure 4.

**Figure 4:**
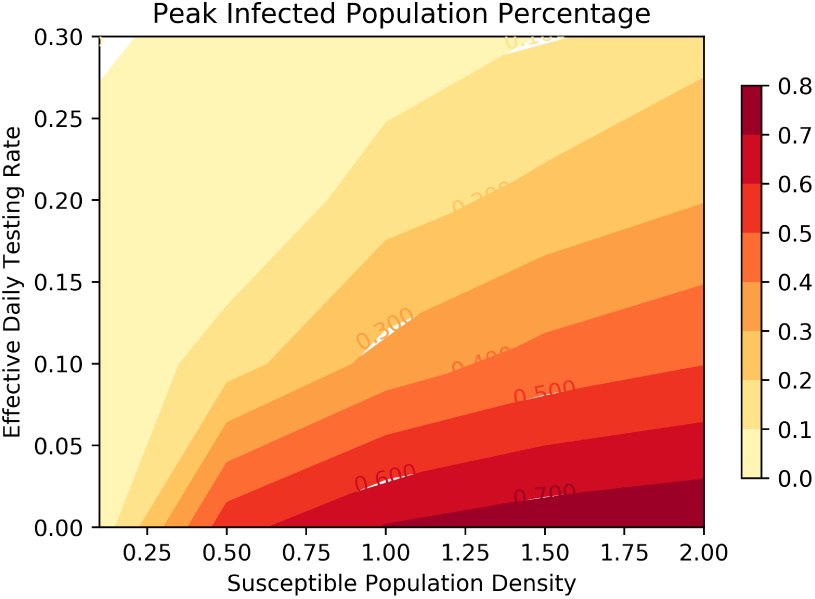
Peak Infection Percentage as a function of *D* and *T*.

## 3.2 Modeling CoVID-19 in Real Regions

The random walk model was used to simulate and visualize the epidemic spread of CoVID-19 in four different regions and countries. Hubei province (China) and South Korea both have experienced a decline in active confirmed cases, while the rate of new active confirmed cases in Spain appears to be decreasing. Cases in Iran continue to increase. Active confirmed cases are calculated by subtracting recovered and dead counts from the number of total confirmed cases. Active confirmed cases are analogous to the combined populations of Infected and Quarantined agents in the model. The simulations are scaled with a linear population scaling factor, and the population density index *D* and testing rate *T* are manually fitted from the rate of active confirmed and removed cases in the real data. The results of these simulations are shown in figures 5 and 6. The parameters used in the simulations are included in table 1.

**Table 1:** Parameters used in Simulations

| Country/Region | $D$ | $T$ | P.S.F. |
| --- | --- | --- | --- |
| Hubei, China | 1.0 | 0.10 | 8 |
| South Korea | 1.5 | 0.20 | 0.8 |
| Spain | 0.8 | 0.12 | 22 |
| Iran | 0.3 | 0.03 | 80 |

**Figure 5:**
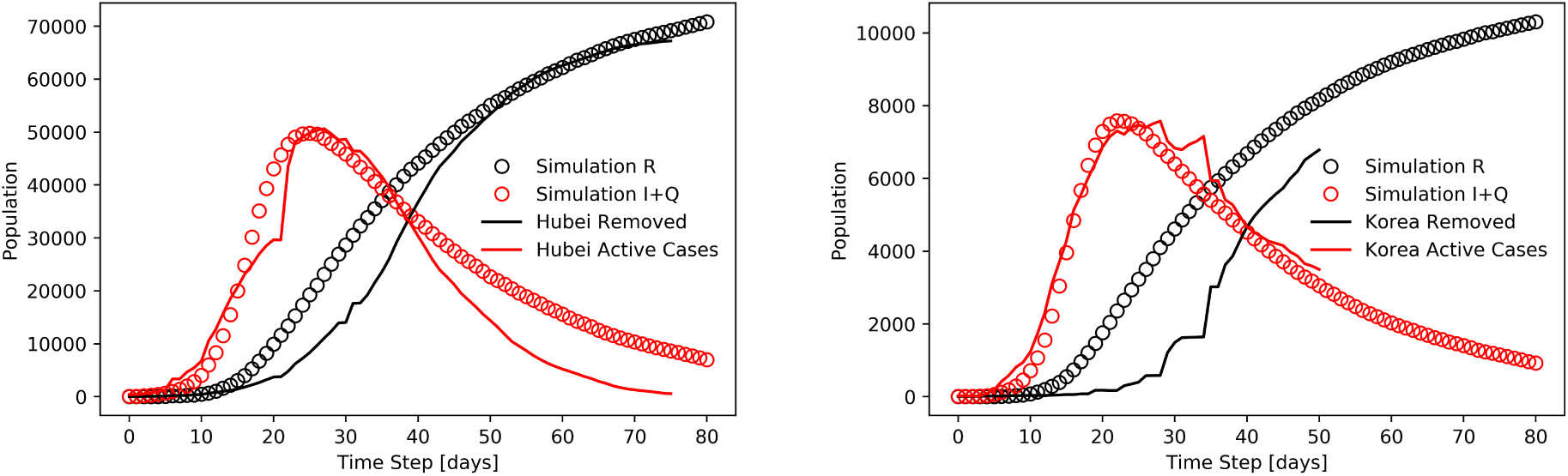
Random Walk Simulation fitted to data [14] from Hubei Province China and South Korea.

**Figure 6:**
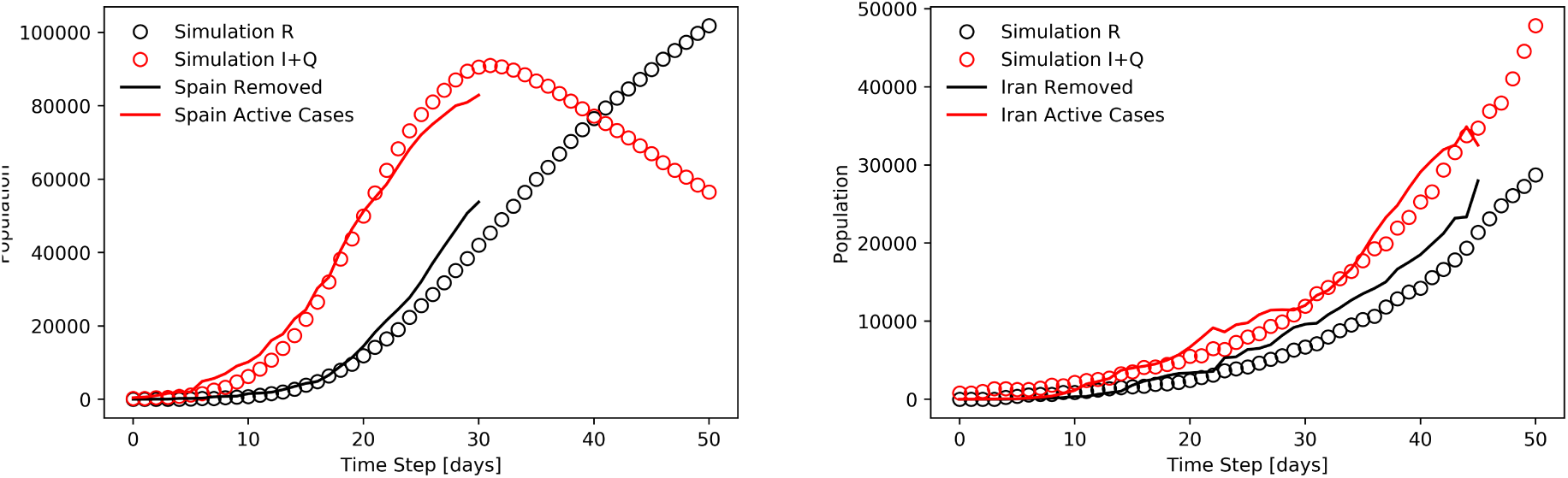
Random Walk Simulation fitted to data [14] from Spain and Iran.

An interactive webpage with epidemic spreading visualizations powered by the model described in this paper can be found at the following address, below.

~~~
pandemic-simulator.mit.edu
~~~

## 4 Discussion

The primary aim of this work was to understand why different regions of the world experience the pandemic spread of CoVID-19 in different ways. By recurrent testing of the population, infected individuals can be identified and quarantined, effectively preventing them from spreading the disease any further. It was observed that through increased rates of testing, an increasing percentage of the population could be saved from experiencing CoVID-19 altogether, to a point where if 40% of the population could be tested daily, any disease having a similar reproduction number would not be able to spread at all. This suggests an opportunity for preventing future viral outbreaks if testing can become a routine procedure that responsible members of the population carry out on themselves and self-report. It was also observed that through increased rates of testing, the peak of confirmed cases could be lowered, relieving the toll of an epidemic on the supply chain, and indirectly human wellbeing.

It was observed that by de-densification of the susceptible population, the peak count of active confirmed infected cases could be reduced and also delayed, to where the rate of infection of the population was decreased, potentially allowing for societal adjustment preceding the time of peak infection. This observation can be an opportunity for actionable advice for urban areas with flexible population densities, such as those with large student populations.

By analyzing how Hubei, China, South Korea, Spain, and Iran have experienced the pandemic spread of CoVID-19, it was observed that countries like South Korea, despite having highly dense susceptible populations, can curb the transmission of the disease in the population by high rates of testing. Along similar lines, it was observed that despite having a less dense population, if recurrent testing rates are low, the potential for an unmitigated outbreak is high as is the case in Iran.

Rather than seek to prescribe a confident policy recommendation or forecast future spreading of the pandemic based on the limited amount of data available currently, this work aims to offer a model for estimating the current spread of CoVID-19 based on demographic information and chosen policies, and present a tool for visualizing the effect that adjusting the variables of population density and testing rate has on a simulated population in a theoretical system.

## Data Availability

All data referred to in manuscript is cited in reference section. The foremost dataset is included in the URL section following, as well as cited in the reference section.

https://github.com/CSSEGISandData/COVID-19/tree/master/csse_covid_19_data/csse_covid_19_time_series

## 5 Acknowledgements

This research was partially funded by the Intelligence Advanced Research Projects Activity (IARPA.) We are grateful to Raj Dandekar, Hyungseok Kim and Wujie Wang for helpful discussions and suggestions.

## References

[1] Ensheng Dong, Hongru Du, and Lauren Gardner. An interactive web-based dashboard to track covid-19 in real time. The Lancet infectious diseases, 2020.

[2] Outbreak of 2019 novel coronavirus (2019-ncov) in wuhan, china, Feb 2020.

[3] Who director-general’s opening remarks at the media briefing on covid-19 - 11 march 2020, Mar 2020.

[4] World Health Organization et al. Coronavirus disease 2019 (covid-19): situation report, 46. 2020.

[5] PT Helo. Dynamic modelling of surge effect and capacity limitation in supply chains. International Journal of Production Research, 38(17):4521–4533, 2000.

[6] Per Bak, Kan Chen, and Chao Tang. A forest-fire model and some thoughts on turbulence. Physics letters A, 147(5-6):297–300, 1990.

[7] Tuen Wai Ng, Gabriel Turinici, and Antoine Danchin. A double epidemic model for the sars propagation. BMC Infectious Diseases, 3(1):19, 2003.

[8] Michael Small, Pengliang Shi, and Chi Kong Tse. Plausible models for propagation of the sars virus. IEICE transactions on fundamentals of electronics, communications and computer sciences, 87(9):2379–2386, 2004.

[9] Boris Shulgin, Lewi Stone, and Zvia Agur. Pulse vaccination strategy in the sir epidemic model. Bulletin of mathematical biology, 60(6):1123–1148, 1998.

[10] Joacim Rocklöv, Henrik Sjödin, and Annelies Wilder-Smith. Covid-19 outbreak on the diamond princess cruise ship: estimating the epidemic potential and effectiveness of public health countermeasures. Journal of Travel Medicine, 2020.

[11] Ying Liu, Albert A Gayle, Annelies Wilder-Smith, and Joacim Rocklöv. The reproductive number of covid-19 is higher compared to sars coronavirus. Journal of travel medicine, 2020.

[12] Biao Tang, Nicola Luigi Bragazzi, Qian Li, Sanyi Tang, Yanni Xiao, and Jianhong Wu. An updated estimation of the risk of transmission of the novel coronavirus (2019-ncov). Infectious disease modelling, 5:248–255, 2020.

[13] Raj Dandekar and George Barbastathis. Quantifying the effect of quarantine control in covid-19 infectious spread using machine learning. medRxiv, 2020.

[14] Hannah Ritchie Max Roser and Esteban Ortiz-Ospina. Coronavirus disease (covid-19) – statistics and research. Our World in Data, 2020. https://ourworldindata.org/coronavirus.

